# The Use of Machine Learning in Occupational Risk Communication for Healthcare Workers – Protocol for scoping review

**DOI:** 10.1101/2024.10.14.24315488

**Authors:** Gabriela Laudares Albuquerque de Oliveira, Clarice Alves Bonow, Itiberê de Oliveira Castellano Rodrigues, Amanda Xavier Geraldo

**Affiliations:** Master in Administration. PhD student in Nursing. Universidade Federal de Pelotas (UFPEL); PhD in Nursing. Universidade Federal de Pelotas (UFPEL); Professor in the area of State Law at the Faculty of Law of the Universidade Federal de Pelotas (UFPEL); Master’s student in Nursing. Universidade Federal de Pelotas (UFPEL)

**Keywords:** Machine Learning, risk communication, occupational health and safety, healthcare workers

## Abstract

**Introduction:** With the development of technology, the use of machine learning (ML), a branch of computer science that aims to transform computers into decision-making agents through algorithms, has grown exponentially. This protocol arises from the need to explore the best practices for applying ML in the communication and management of occupational risks for healthcare workers.

**Methods and analysis:** This scoping review protocol^1^ details a search to be conducted in the academic databases Public Medical Literature Analysis and Retrieval System Online (PUBMED), through the Virtual Health Library (BVS): Medical Literature Analysis and Retrieval System (MEDLINE), Latin American and Caribbean Literature in Health Sciences (LILACS), West Pacific (WPRIM), Nursing Database (BDENF) and Scientific Electronic Library Online (SciELO), SCOPUS, Web of Science, IEEE Xplore Digital Library and Excerpta Medica Database (EMBASE). This scoping review protocol outlines the objectives, methods, and timeline for a review that will explore and map the existing scientific evidence and knowledge on the use of machine learning (ML) in risk communication for healthcare workers. This protocol follows the PRISMA-ScR^2^ and JBI guidelines^3^ for conducting scoping reviews. The guiding question of the review is: How is machine learning used in risk communication for healthcare workers? The search will use PCC (Population, Concept, Context) terms and the specific descriptors defined by each database. The narrative synthesis will describe the main themes and findings of the review.

The results of this scoping review will be disseminated through publication in an international peer-reviewed scientific journal.

**Ethics and dissemination:** Ethical approval is not required; data will rely on published articles. Findings will be published open access in an international peer-reviewed journal.

**Strengths and limitations of this study:** *Strengths:* The study allows for comprehensive mapping of existing evidence on ML in occupational risk communication. The methodology follows PRISMA-ScR and JBI guidelines, ensuring transparency and replicability. The research employs a broad search strategy across multiple databases to capture relevant studies.

*Limitations:* The accuracy of ML models is dependent on the quality of the data used. The implementation of ML in healthcare requires careful evaluation of ethical, legal, and privacy issues. **Registration details OSF Registries:** The protocol for this review was registered in the Open Science Framework under DOI 10.17605/OSF.IO/92SK4 (available at https://osf.io/92SK4).

## INTRODUCTION

The International Labour Organization^4^ estimates that approximately 2.3 million workers die annually due to work-related accidents or illnesses, which corresponds to more than 6,000 deaths per day. In Brazil, according to the Occupational Health and Safety Observatory, over 26,000 deaths and 6,804,060 Work Accident Notifications (CAT) were recorded between 2012 and 2022, with an average of approximately 618,000 accidents reported annually, while underreporting is estimated at nearly 19%. The observatory highlights priority areas, with Nursing Technicians being the occupation most affected by recorded Work Accidents (more than 313,000 between 2012 and 2022), followed by Nurses and Nursing Assistants, with 8,673 and 5,472 cases, respectively. These figures underscore the severity of this global issue^5^.

Therefore, the shortage of nursing professionals is recognized as a global health emergency^6^, and unsatisfactory working conditions are directly linked to absenteeism and turnover^7 8 9^.

Health care workers face a variety of occupational risks, ranging from exposure to biological and chemical risks, physical risks, ergonomic risks and psychosocial risks such as shift work and workplace violence^7^. In this context, risk communication plays a crucial role in informing, educating, and engaging workers about hazards and available protective measures^8 10^.

The integration of artificial intelligence (AI) and Machine Learning in risk communication can yield significant benefits by enabling the analysis of large volumes of data to identify patterns and predict potential risks^9^. These technologies have the potential to enhance the safety of healthcare professionals, although their implementation requires ethical, legal, and privacy considerations^12 13 14^.

Previous studies have shown that the application of Machine Learning (ML) algorithms, such as neural networks, not only increases the accuracy of risk prediction, but also facilitates the customization of communication strategies to meet the needs of different audiences. For instance, a study conducted with teachers found that it is possible to effectively predict cases of absenteeism due to morbidity, yielding promising results. The algorithm with the best predictive performance, according to standard ML evaluation metrics, was artificial neural networks, achieving an area under the curve of 0.79, with an accuracy of 71.52%, sensitivity of 72.86%, and specificity of 70.52%^15^.

In Brazil, although there are no specific AI strategies for risk communication in the healthcare sector yet, it is crucial for this to be an area of interest to ensure the safety and well-being of workers and to anticipate safer practices in the face of imminent risks^15^.

A preliminary search was conducted in The Cochrane Library, Open Science Framework, Database of Abstracts of Reviews of Effects (DARE), and the International Prospective Register of Systematic Reviews (PROSPERO), and no reviews or studies similar to this research were found.

Through this approach, we intend to investigate how artificial intelligence (AI) and machine learning can contribute to the prevention of occupational accidents and diseases in the healthcare sector. By analyzing relevant studies and applications, we seek to identify effective strategies to protect healthcare professionals from various risks, such as biological, chemical, ergonomic, psychosocial, and physical.

Therefore, this review aims to explore the potential of AI in risk communication for healthcare professionals, analyzing relevant studies and applications to guide future applications in Brazil.

## METHODS AND ANALYSIS

The choice of a scoping review to address the topic “Use of Machine Learning in risk communication for healthcare workers” is justified by the need to explore and map the available scientific evidence and existing knowledge in this specific area. This protocol and review follow the guidelines and recommendations^1^ of the Preferred Reporting Items for Systematic reviews and Meta-Analyses extension for Scoping Reviews (PRISMA-ScR) checklist^2^ and JBI (Joanna Briggs Institute)^3^. The scoping review will meet the premises of this research, as it does not intend to assess the quality of available evidence but rather to obtain a representation of the findings.

In the specific case of the scoping review on the use of Machine Learning in occupational risk communication for healthcare workers, this methodology will allow for a comprehensive and systematic search for different types of scientific articles. By adhering to the guidelines and recommendations of Prisma^2^ or JBI^3^, the scoping review will ensure transparency, rigor, and replicability in all stages of the process, from formulating the research question to analyzing and synthesizing the findings. This will help provide a comprehensive and up-to-date overview of the use of Machine Learning in occupational risk communication for healthcare workers, contributing to informed decision-making and the development of effective interventions in this area.

The review process^1^ will follow the steps recommended by PRISMA-ScR^2^, including Introduction, Review Question, Inclusion Criteria (Participants, Concept, Context), Types of Sources, Methods, Search Strategy, Study/Source of Evidence Selection, Data Extraction and Data Analysis and Presentation. A version of PRISMA-ScR as it applies to this protocol document has been added in online supplemental appendix I.

### Review question

The research question was developed through alignment with the core elements/PCC framework, P = Population/ Participants, C = Concept, C = Context.

The PCC methodology is a widely used approach in scoping reviews to outline the characteristics of the population of interest, the relevant concepts or phenomena, and the context in which they occur, as recommended in the JBI Manual for Scoping Reviews^3^. In this study, we applied the PCC methodology to identify key elements related to the communication of occupational risks for workers exposed to hazardous work environments.

P - Population: Healthcare workers exposed to occupational risks.

C - Concept: Machine Learning is the conceptual technique to be explored in this scoping review.

C - Context: Occupational risk communication. The aim is to examine how effective communication of occupational risks can contribute to accident and work-related illness prevention, as well as to identify barriers and challenges encountered in this process.

With this mnemonic combination, the following guiding question was defined:

How has the use of Machine Learning been applied in risk communication for healthcare workers?

This question provides a clear direction for investigating the application of artificial intelligence in risk communication for healthcare workers and evaluating its outcome.

Before conducting this study, searches were conducted on The Cochane Library, Open Science Framework, Database of Abstracts of Reviews of Effects (DARE), and the International Prospective Register of Systematic Reviews (PROSPERO) websites to locate similar reviews or studies and prevent study duplication. No similar studies were found and this scoping review protocol was registered in the Open Science Framework under a protocol: DOI 10.17605/OSF.IO/92SK4 (available at https://osf.io/92SK4).

Google Scholar searches^13 14 16 17 18^ related to the topic were conducted using the PCC keywords to identify the most recurrent descriptors and keywords. These descriptors and keywords were then listed in a spreadsheet according to Table 1 to initiate searches in PubMed.

**Table 1:**
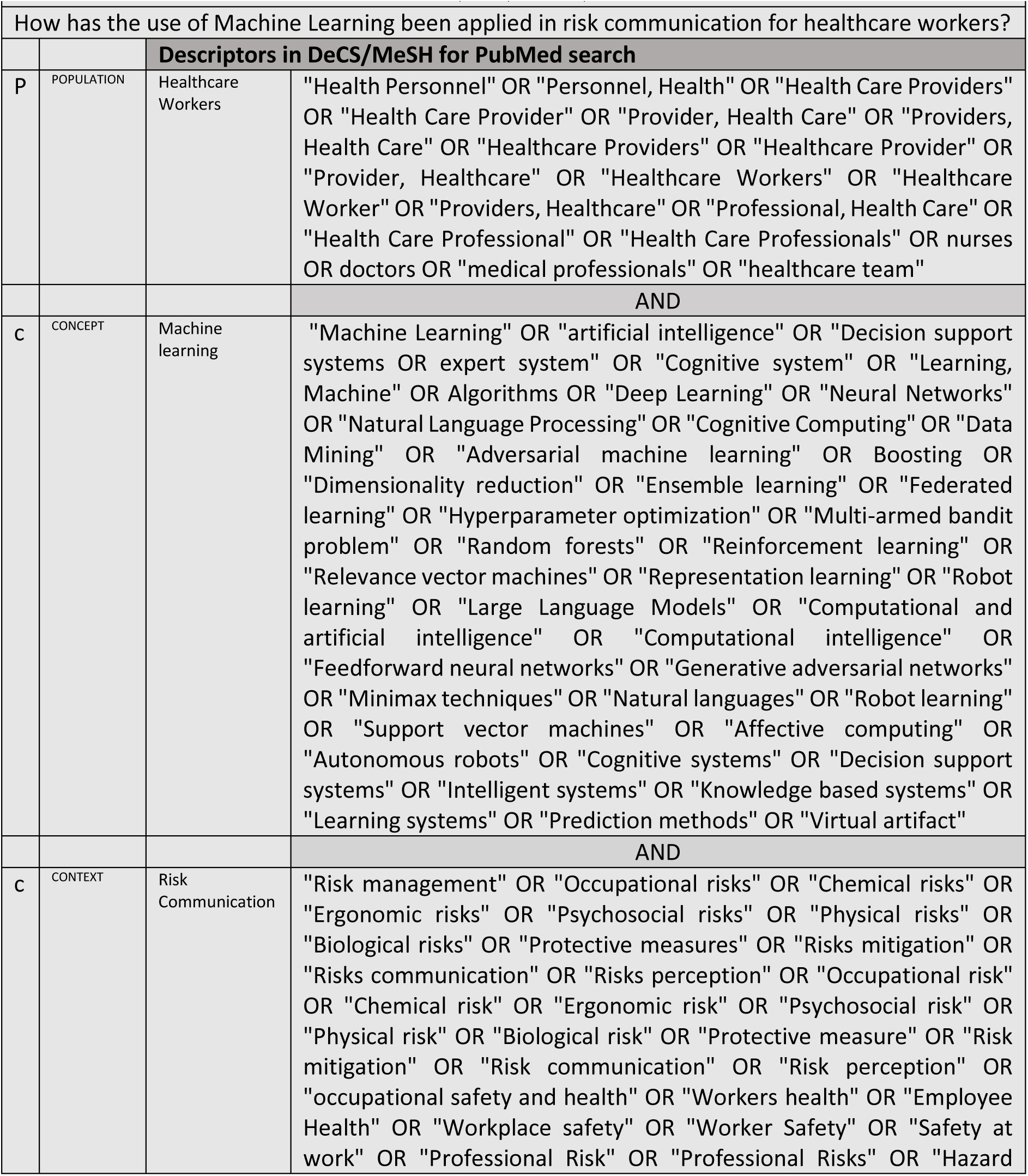
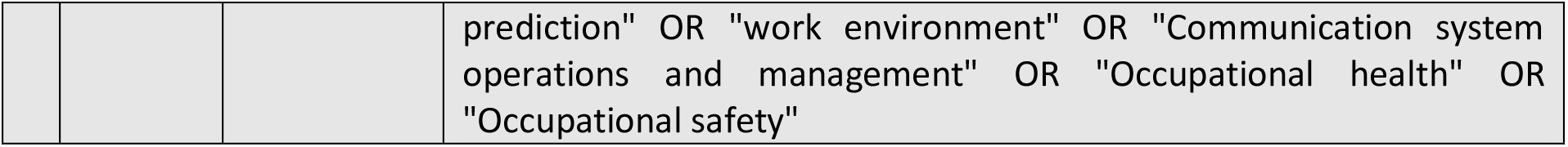
Searches in PubMed - Pelotas, RS, Brazil, 2024.

### Types of sources and inclusion and exclusion criteria

Inclusion and exclusion criteria will follow the dimensions outlined by the elements of the acronym PCC, with the Population (P) being healthcare workers, nurses, physicians, thus studies dealing with other populations (such as patients) will be excluded. In the Concept (C) element, all studies presenting the use of Artificial Intelligence, especially Machine Learning, will be included, while studies presenting other tools in their concept will be excluded. For Context (C), studies presenting communication and strategies for occupational risk management will be considered, while studies focused on other communication contexts (such as clinical decision support, patient safety, among others) will be excluded.

The criterion defined for selecting the databases used PUBMED, MEDLINE, LILACS, WPRIM, BDENF, SciELO, SCOPUS, Web of Science, EMBASE, IEEE Xplore, will be the availability to search for articles through search engines with support for descriptors and boolean operators, as they are updated databases. Studies published in all languages will be selected. After this step, the snowballing technique will be used to review the references of all included articles to identify other studies that may also meet the selection criteria.

The general inclusion criteria for articles will be: articles published in the last 5 years, reflecting advancements in machine learning (ML) driven by global regulatory milestones like the European Union's GDPR (2018). Such regulations have set ethical and transparency standards for data use, promoting safer ML techniques aligned with data protection, especially in occupational health and safety contexts19.

The general exclusion criteria will be: incomplete articles, articles not available in full, and grey literature (theses and dissertations, conference proceedings, reports, government documents, among others). It is worth noting that grey literature was not prioritized given its various dissemination interests, in addition to the scientific one that corresponds to the unitary focus of this study. Even though theses and dissertations, for example, are in academic and therefore scientific contexts, they were not included, as it is understood that the level of academic and scientific equity is achieved in the publication of articles in peer-reviewed journals.

### Search Strategy

The electronic search will be conducted between May and October 2024, using Health Science Descriptors (DeCS) in English, or Medical Subject Headings (MeSH), terms found in the IEEE Thesaurus and EMTREE Thesaurus (Embase). These terms will be used to search the following databases: Public Medical Literature Analysis and Retrieval System Online (PUBMED), through the Virtual Health Library (VHL): MEDLINE, Latin American and Caribbean Health Sciences Literature (LILACS), WPRIM, BDENF, and Scientific Electronic Library Online (SciELO), SCOPUS, Web of Science, IEEE Xplore, and EMBASE. The search strategy was adjusted as per the requirements of the different databases as indicated in the search strategy presented in the online supplementary appendix II.

As per Table 1. After each descriptor/keyword/search strategy search, titles of articles found will be reviewed, focusing on terms related to the research topic, following the PCC strategy. When the title and abstract are unclear, introduction and conclusion will be read.

### Study Selection

The description and summarization of data will be conducted by two independent reviewers for reviewing titles, abstracts, and keywords for studies meeting inclusion criteria. In case of any disagreement, a third reviewer will be called upon to analyze and decide on the inclusion or exclusion of articles.

Zotero and Rayyan managers will be used to identify duplicate articles and organize eligible or non-eligible studies.

### Data Extraction

For information mapping, data collection will be conducted using an instrument adapted from the JBI^2^ form, developed by the authors in Microsoft Excel to record relevant study categories, as shown in Table 2.

**Table 2:**
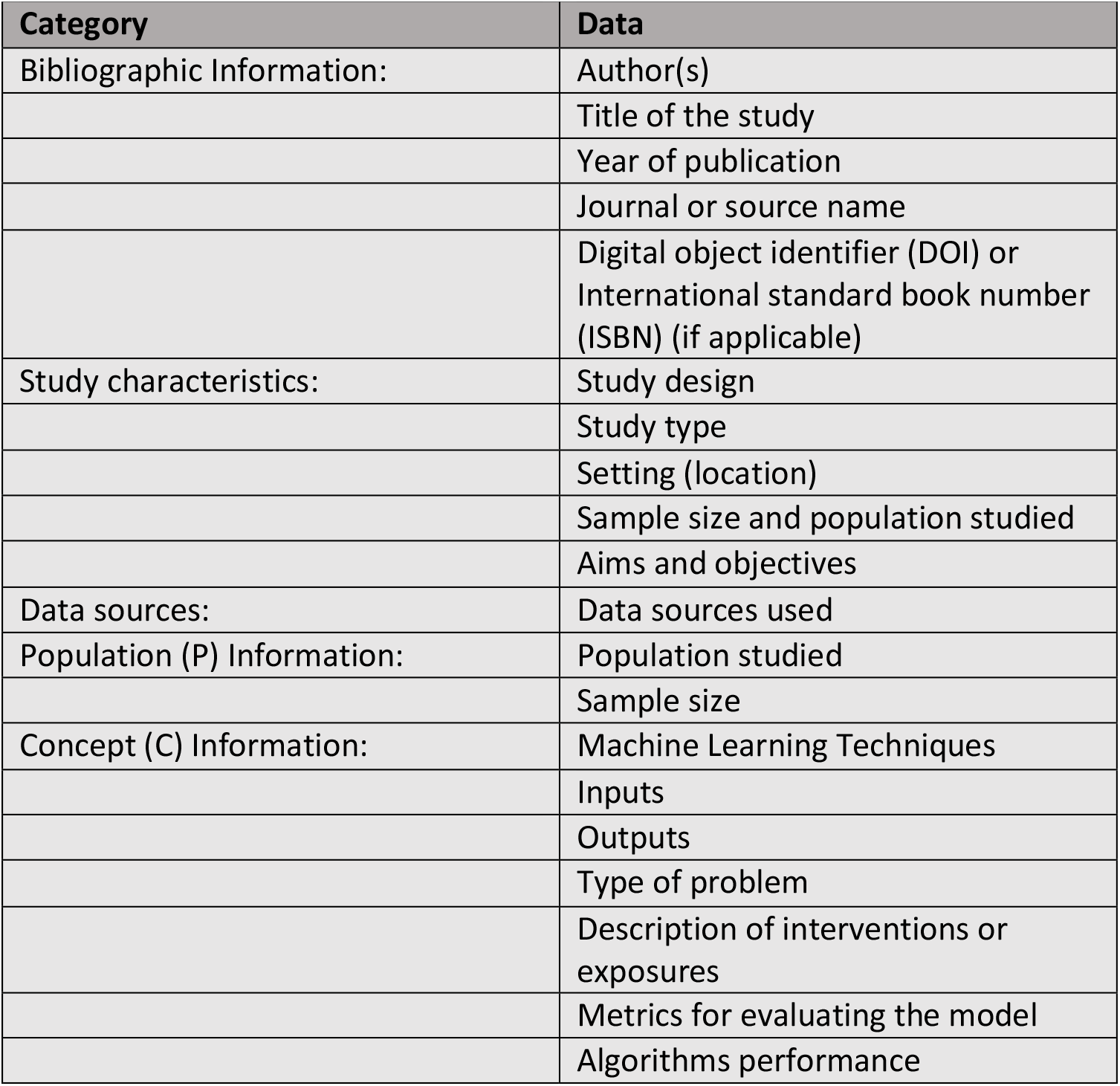
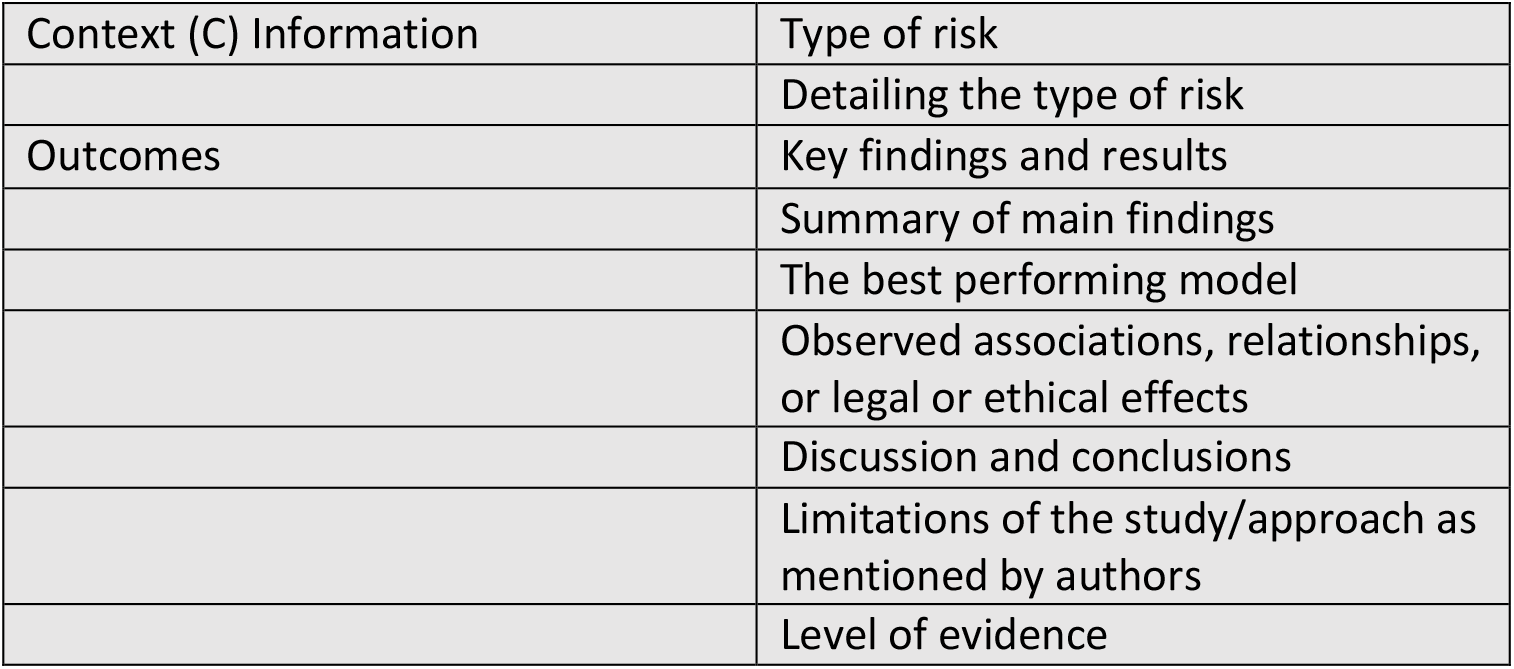
Data items to be collected.

| Table 2 Data items to be collected |  |
| --- | --- |
| Category | Data |
| Bibliographic Information: | Author(s) |
|  | Title of the study |
|  | Year of publication |
|  | Journal or source name |
|  | Digital object identifier (DOI) or International standard book number (ISBN) (if applicable) |
| Study characteristics: | Study design |
|  | Study type |
|  | Setting (location) |
|  | Sample size and population studied |
|  | Aims and objectives |
| Data sources: | Data sources used |
| Population (P) Information: | Population studied |
|  | Sample size |
| Concept (C) Information: | Machine Learning Techniques |
|  | Inputs |
|  | Outputs |
|  | Type of problem |
|  | Description of interventions or exposures |
|  | Metrics for evaluating the model |
|  | Algorithms performance |
| Context (C) Information | Type of risk |
|  | Detailing the type of risk |
| Outcomes | Key findings and results |
|  | Summary of main findings |
|  | The best performing model |
|  | Observed associations, relationships, or legal or ethical effects |
|  | Discussion and conclusions |
|  | Limitations of the study/approach as mentioned by authors |
|  | Level of evidence |

### Planned dates

The study is planned to begin in May 2024 and end in December 2024.

## Supporting information

PRISMA ScR Checklist

Search Strategy

## Data Availability

Data availability statement
No data are available.

https://drive.google.com/file/d/1qWIUThBJon6sbclvZapx7QFox2EUGYgI/view?usp=drive_link

https://drive.google.com/file/d/1YGXJqx-gJnr80vkteLamCfXUKRLy78w6/view?usp=drive_link

## Acknowledgements

This review is part of a PhD trajectory of Oliveira, GLA.

## Author contributions

GLAO, AXG, CAB, and IOCR contributed to the development of this study protocol. CAB and GLAO proposed the study, which was then developed by GLAO and AXG with the help of all the authors. GLAO, AXG, CAB, and IOCR discussed the results of the exploratory search and prepared the supporting theoretical framework. CAB and IOCR provided specialized knowledge in Risk Communication and legal aspects of Artificial Intelligence use, respectively. GLAO identified the search terms and established the search strategy together with CAB and the university library. The scoping review planning will follow the following strategy: GLAO and AXG will be responsible for data collection and will read the titles and abstracts of the articles found after each search for descriptors/keywords, looking for words related to the research topic, according to the PCC strategy. When the title and abstract are not clear, the introduction and conclusion or the entire study will be read if necessary, and if there is any disagreement, CAB will be called to analyze and decide whether or not to include the articles. After this step, the articles and the work will be divided among the authors for screening and secondary analysis. GLAO will prepare the first drafts of the manuscript. The final manuscript will be prepared through interactive refinement and group discussions between GLAO, AXG, CAB, and IOCR.

## Funding

GLAB is supported by a doctoral scholarship [[141906/2023-5] from the National Council for Scientific and Technological Development (CNPq).

## Competing interests

None declared.

## Patient and public involvement

Patients and/or the public were not involved in the design, or conduct, or reporting or dissemination plans of this research.

## Patient consent for publication

Not applicable.

## Ethics and dissemination

This review does not require ethical approval. Our dissemination strategy includes publishing in peer-reviewed journals, presenting at conferences, and sharing findings with relevant stakeholders.

## Data availability statement

No data are available.

## Notes

### Competing Interest Statement

The authors have declared no competing interest.

### Summary of Updates

Response to Reviewer Comments - GABRIELA LAUDARES ALBUQUERQUE DE OLIVEIRA - APPENDIX

