## Supplementary material for "The Use of Machine Learning in Occupational Risk Communication for Healthcare Workers – Protocol for scoping review": Search Strategy

### Appendix II: Search strategies

**Search Strategy for PubMed (11 October 2024)**

| Search number | Query | Results | Time |
|---|---|---|---|
| #4 | #1 AND #2 AND #3 | 979 | 14:08 |
| #3 | "Risk management" OR "Occupational risks" OR "Chemical risks" OR "Ergonomic risks" OR "Psychosocial risks" OR "Physical risks" OR "Biological risks" OR "Protective measures" OR "Risks mitigation" OR "Risks communication" OR "Risks perception" OR "Occupational risk" OR "Chemical risk" OR "Ergonomic risk" OR "Psychosocial risk" OR "Physical risk" OR "Biological risk" OR "Protective measure" OR "Risk mitigation" OR "Risk communication" OR "Risk perception" OR "occupational safety and health" OR "Workers health" OR "Employee Health" OR "Workplace safety" OR "Worker Safety" OR "Safety at work" OR "Professional Risk" OR "Professional Risks" OR "Hazard prediction" OR "work environment" OR "Communication system operations and management" OR "Occupational health" OR "Occupational safety" | 264.230 | 14:08 |
| #2 | "Machine Learning" OR "artificial intelligence" OR "Decision support systems" OR "expert system" OR "Cognitive system" OR "Learning, Machine" OR Algorithms OR "Deep Learning" OR "Neural Networks" OR "Natural Language Processing" OR "Cognitive Computing" OR "Data Mining" OR "Adversarial machine learning" OR Boosting OR "Dimensionality reduction" OR "Ensemble learning" OR "Federated learning" OR "Hyperparameter optimization" OR "Multi-armed bandit problem" OR "Random forests" OR "Reinforcement learning" OR "Relevance vector machines" OR "Representation learning" OR "Robot learning" OR "Large Language Models" OR "Computational and artificial intelligence" OR "Computational intelligence" OR "Feedforward neural networks" OR "Generative adversarial networks" OR "Minimax techniques" OR "Natural languages" OR "Robot learning" OR "Support vector machines" OR "Affective computing" OR "Autonomous robots" OR "Cognitive systems" OR "Decision support systems" OR "Intelligent systems" OR "Knowledge based systems" OR "Learning systems" OR "Prediction methods" OR "Virtual artifact" | 96.137 | 14:08 |
| #1 | "Health Personnel" OR "Personnel, Health" OR "Health Care Providers" OR "Health Care Provider" OR "Provider, Health Care" OR "Providers, Health Care" OR "Healthcare Providers" OR "Healthcare Provider" OR "Provider, Healthcare" OR "Healthcare Workers" OR "Healthcare Worker" OR "Providers, Healthcare" OR "Professional, Health Care" OR "Health Care Professional" OR "Health Care Professionals" OR nurses OR doctors OR "medical professionals" OR "healthcare team" | 1.430.209 | 14:07 |

**Search strategies 11 October 2024**
**Search Strategy for Medline/ BVS (11 October 2024)**

| Query | Results | Time |
|---|---|---|
| ("Health Personnel" OR "Personnel, Health" OR "Health Care Providers" OR "Health Care Provider" OR "Provider, Health Care" OR "Providers, Health Care" OR "Healthcare Providers" OR "Healthcare Provider" OR "Provider, Healthcare" OR "Healthcare Workers" OR "Healthcare Worker" OR "Providers, Healthcare" OR "Professional, Health Care" OR "Health Care Professional" OR "Health Care Professionals" OR nurses OR doctors OR "medical professionals" OR "healthcare team") AND ( "Machine Learning" OR "artificial intelligence" OR "Decision support systems" OR "expert system" OR "Cognitive system" OR "Learning, Machine" OR algorithms OR "Deep Learning" OR "Neural Networks" OR "Natural Language Processing" OR "Cognitive Computing" OR "Data Mining" OR "Adversarial machine learning" OR boosting OR "Dimensionality reduction" OR "Ensemble learning" OR "Federated learning" OR "Hyperparameter optimization" OR "Multi-armed bandit problem" OR "Random forests" OR "Reinforcement learning" OR "Relevance vector machines" OR "Representation learning" OR "Robot learning" OR "Large Language Models" OR "Computational and artificial intelligence" OR "Computational intelligence" OR "Feedforward neural networks" OR "Generative adversarial networks" OR "Minimax techniques" OR "Natural languages" OR "Robot learning" OR "Support vector machines" OR "Affective computing" OR "Autonomous robots" OR "Cognitive systems" OR "Decision support systems" OR "Intelligent systems" OR "Knowledge based systems" OR "Learning systems" OR "Prediction methods" OR "Virtual artifact") AND ("Risk management" OR "Occupational risks" OR "Chemical risks" OR "Ergonomic risks" OR "Psychosocial risks" OR "Physical risks" OR "Biological risks" OR "Protective measures" OR "Risks mitigation" OR "Risks communication" OR "Risks perception" OR "Occupational risk" OR "Chemical risk" OR "Ergonomic risk" OR "Psychosocial risk" OR "Physical risk" OR "Biological risk" OR "Protective measure" OR "Risk mitigation" OR "Risk communication" OR "Risk perception" OR "occupational safety and health" OR "Workers health" OR "Employee Health" OR "Workplace safety" OR "Worker Safety" OR "Safety at work" OR "Professional Risk" OR "Professional Risks" OR "Hazard prediction" OR "work environment" OR "Communication system operations and management" OR "Occupational health" OR "Occupational safety") AND ( db:("MEDLINE")) | 203 | 15:37 |

**Search strategies 11 October 2024**
**Search Strategy for Lilacs/ BVS (11 October 2024)**

| Query | Results | Time |
|---|---|---|
| ("Health Personnel" OR "Personnel, Health" OR "Health Care Providers" OR "Health Care Provider" OR "Provider, Health Care" OR "Providers, Health Care" OR "Healthcare Providers" OR "Healthcare Provider" OR "Provider, Healthcare" OR "Healthcare Workers" OR "Healthcare Worker" OR "Providers, Healthcare" OR "Professional, Health Care" OR "Health Care Professional" OR "Health Care Professionals" OR nurses OR doctors OR "medical professionals" OR "healthcare team") AND ( "Machine Learning" OR "artificial intelligence" OR "Decision support systems" OR "expert system" OR ("Health Personnel" OR "Personnel, Health" OR "Health Care Providers" OR "Health Care Provider" OR "Provider, Health Care" OR "Providers, Health Care" OR "Healthcare Providers" OR "Healthcare Provider" OR "Provider, Healthcare" OR "Healthcare Workers" OR "Healthcare Worker" OR "Providers, Healthcare" OR "Professional, Health Care" OR "Health Care Professional" OR "Health Care Professionals" OR nurses OR doctors OR "medical professionals" OR "healthcare team") AND ( "Machine Learning" OR "artificial intelligence" OR "Decision support systems" OR "expert system" OR "Cognitive system" OR "Learning, Machine" OR algorithms OR "Deep Learning" OR "Neural Networks" OR "Natural Language Processing" OR "Cognitive Computing" OR "Data Mining" OR "Adversarial machine learning" OR boosting OR "Dimensionality reduction" OR "Ensemble learning" OR "Federated learning" OR "Hyperparameter optimization" OR "Multi-armed bandit problem" OR "Random forests" OR "Reinforcement learning" OR "Relevance vector machines" OR "Representation learning" OR "Robot learning" OR "Large Language Models" OR "Computational and artificial intelligence" OR "Computational intelligence" OR "Feedforward neural networks" OR "Generative adversarial networks" OR "Minimax techniques" OR "Natural languages" OR "Robot learning" OR "Support vector machines" OR "Affective computing" OR "Autonomous robots" OR "Cognitive systems" OR "Decision support systems" OR "Intelligent systems" OR "Knowledge based systems" OR "Learning systems" OR "Prediction methods" OR "Virtual artifact") AND ("Risk management" OR "Occupational risks" OR "Chemical risks" OR "Ergonomic risks" OR "Psychosocial risks" OR "Physical risks" OR "Biological risks" OR "Protective measures" OR "Risks mitigation" OR "Risks communication" OR "Risks perception" OR "Occupational risk" OR "Chemical risk" OR "Ergonomic risk" OR "Psychosocial risk" OR "Physical risk" OR "Biological risk" OR "Protective measure" OR "Risk mitigation" OR "Risk communication" OR "Risk perception" OR "occupational safety and health" OR "Workers health" OR "Employee Health" OR "Workplace safety" OR "Worker Safety" OR "Safety at work" OR "Professional Risk" OR "Professional Risks" OR "Hazard prediction" OR "work environment" OR "Communication system operations and management" OR "Occupational health" OR "Occupational safety") AND ( db:("LILACS")) | 6 | 15:39 |

**Search strategies 11 October 2024**
**Search Strategy for BDENF/ BVS (11 October 2024)**

| Query | Results | Time |
|---|---|---|
| ("Health Personnel" OR "Personnel, Health" OR "Health Care Providers" OR "Health Care Provider" OR "Provider, Health Care" OR "Providers, Health Care" OR "Healthcare Providers" OR "Healthcare Provider" OR "Provider, Healthcare" OR "Healthcare Workers" OR "Healthcare Worker" OR "Providers, Healthcare" OR "Professional, Health Care" OR "Health Care Professional" OR "Health Care Professionals" OR nurses OR doctors OR "medical professionals" OR "healthcare team") AND ( "Machine Learning" OR "artificial intelligence" OR "Decision support systems" OR "expert system" OR "Cognitive system" OR "Learning, Machine" OR algorithms OR "Deep Learning" OR "Neural Networks" OR "Natural Language Processing" OR "Cognitive Computing" OR "Data Mining" OR "Adversarial machine learning" OR boosting OR "Dimensionality reduction" OR "Ensemble learning" OR "Federated learning" OR "Hyperparameter optimization" OR "Multi-armed bandit problem" OR "Random forests" OR "Reinforcement learning" OR "Relevance vector machines" OR "Representation learning" OR "Robot learning" OR "Large Language Models" OR "Computational and artificial intelligence" OR "Computational intelligence" OR "Feedforward neural networks" OR "Generative adversarial networks" OR "Minimax techniques" OR "Natural languages" OR "Robot learning" OR "Support vector machines" OR "Affective computing" OR "Autonomous robots" OR "Cognitive systems" OR "Decision support systems" OR "Intelligent systems" OR "Knowledge based systems" OR "Learning systems" OR "Prediction methods" OR "Virtual artifact") AND ("Risk management" OR "Occupational risks" OR "Chemical risks" OR "Ergonomic risks" OR "Psychosocial risks" OR "Physical risks" OR "Biological risks" OR "Protective measures" OR "Risks mitigation" OR "Risks communication" OR "Risks perception" OR "Occupational risk" OR "Chemical risk" OR "Ergonomic risk" OR "Psychosocial risk" OR "Physical risk" OR "Biological risk" OR "Protective measure" OR "Risk mitigation" OR "Risk communication" OR "Risk perception" OR "occupational safety and health" OR "Workers health" OR "Employee Health" OR "Workplace safety" OR "Worker Safety" OR "Safety at work" OR "Professional Risk" OR "Professional Risks" OR "Hazard prediction" OR "work environment" OR "Communication system operations and management" OR "Occupational health" OR "Occupational safety") AND ( db:("BDENF")) | 1 | 15:40 |

**Search strategies 11 October 2024**

**Search Strategy for WPRIM/ BVS (11 October 2024)**

| Query | Results | Time |
|---|---|---|
| ("Health Personnel" OR "Personnel, Health" OR "Health Care Providers" OR "Health Care Provider" OR "Provider, Health Care" OR "Providers, Health Care" OR "Healthcare Providers" OR "Healthcare Provider" OR "Provider, Healthcare" OR "Healthcare Workers" OR "Healthcare Worker" OR "Providers, Healthcare" OR "Professional, Health Care" OR "Health Care Professional" OR "Health Care Professionals" OR nurses OR doctors OR "medical professionals" OR "healthcare team") AND ( "Machine Learning" OR "artificial intelligence" OR "Decision support systems" OR "expert system" OR "Cognitive system" OR "Learning, Machine" OR algorithms OR "Deep Learning" OR "Neural Networks" OR "Natural Language Processing" OR "Cognitive Computing" OR "Data Mining" OR "Adversarial machine learning" OR boosting OR "Dimensionality reduction" OR "Ensemble learning" OR "Federated learning" OR "Hyperparameter optimization" OR "Multi-armed bandit problem" OR "Random forests" OR "Reinforcement learning" OR "Relevance vector machines" OR "Representation learning" OR "Robot learning" OR "Large Language Models" OR "Computational and artificial intelligence" OR ("Health Personnel" OR "Personnel, Health" OR "Health Care Providers" OR "Health Care Provider" OR "Provider, Health Care" OR "Providers, Health Care" OR "Healthcare Providers" OR "Healthcare Provider" OR "Provider, Healthcare" OR "Healthcare Workers" OR "Healthcare Worker" OR "Providers, Healthcare" OR "Professional, Health Care" OR "Health Care Professional" OR "Health Care Professionals" OR nurses OR doctors OR "medical professionals" OR "healthcare team") AND ( "Machine Learning" OR "artificial intelligence" OR "Decision support systems" OR "expert system" OR "Cognitive system" OR "Learning, Machine" OR algorithms OR "Deep Learning" OR "Neural Networks" OR "Natural Language Processing" OR "Cognitive Computing" OR "Data Mining" OR "Adversarial machine learning" OR boosting OR "Dimensionality reduction" OR "Ensemble learning" OR "Federated learning" OR "Hyperparameter optimization" OR "Multi-armed bandit problem" OR "Random forests" OR "Reinforcement learning" OR "Relevance vector machines" OR "Representation learning" OR "Robot learning" OR "Large Language Models" OR "Computational and artificial intelligence" OR "Computational intelligence" OR "Feedforward neural networks" OR "Generative adversarial networks" OR "Minimax techniques" OR "Natural languages" OR "Robot learning" OR "Support vector machines" OR "Affective computing" OR "Autonomous robots" OR "Cognitive systems" OR "Decision support systems" OR "Intelligent systems" OR "Knowledge based systems" OR "Learning systems" OR "Prediction methods" OR "Virtual artifact") AND ("Risk management" OR "Occupational risks" OR "Chemical risks" OR "Ergonomic risks" OR "Psychosocial risks" OR "Physical risks" OR "Biological risks" OR "Protective measures" OR "Risks mitigation" OR "Risks communication" OR "Risks perception" OR "Occupational risk" OR "Chemical risk" OR "Ergonomic risk" OR "Psychosocial risk" OR "Physical risk" OR "Biological risk" OR "Protective measure" OR "Risk mitigation" OR "Risk communication" OR "Risk perception" OR "occupational safety and health" OR "Workers health" OR "Employee Health" OR "Workplace safety" OR "Worker Safety" OR "Safety at work" OR "Professional Risk" OR "Professional Risks" OR "Hazard prediction" OR "work environment" OR "Communication system operations and management" OR "Occupational health" OR "Occupational safety") AND ( db:("WPRIM")) | 2 | 15:55 |

**Search strategies 11 October 2024**

**Search Strategy for SCIELO (11 October 2024)**

| Query | Results | Time |
|---|---|---|
| ("Health Personnel" OR "Personnel, Health" OR "Health Care Providers" OR "Health Care Provider" OR "Provider, Health Care" OR "Providers, Health Care" OR "Healthcare Providers" OR "Healthcare Provider" OR "Provider, Healthcare" OR "Healthcare Workers" OR "Healthcare Worker" OR "Providers, Healthcare" OR "Professional, Health Care" OR "Health Care Professional" OR "Health Care Professionals" OR nurses OR doctors OR "medical professionals" OR "healthcare team") AND ( "Machine Learning" OR "artificial intelligence" OR "Decision support systems" OR "expert system" OR "Cognitive system" OR "Learning, Machine" OR Algorithms OR "Deep Learning" OR "Neural Networks" OR "Natural Language Processing" OR "Cognitive Computing" OR "Data Mining" OR "Adversarial machine learning" OR Boosting OR "Dimensionality reduction" OR "Ensemble learning" OR "Federated learning" OR "Hyperparameter optimization" OR "Multi-armed bandit problem" OR "Random forests" OR "Reinforcement learning" OR "Relevance vector machines" OR "Representation learning" OR "Robot learning" OR "Large Language Models" OR "Computational and artificial intelligence" OR "Computational intelligence" OR "Feedforward neural networks" OR "Generative adversarial networks" OR "Minimax techniques" OR "Natural languages" OR "Robot learning" OR "Support vector machines" OR "Affective computing" OR "Autonomous robots" OR "Cognitive systems" OR "Decision support systems" OR "Intelligent systems" OR "Knowledge based systems" OR "Learning systems" OR "Prediction methods" OR "Virtual artifact") AND ("Risk management" OR "Occupational risks" OR "Chemical risks" OR "Ergonomic risks" OR "Psychosocial risks" OR "Physical risks" OR "Biological risks" OR "Protective measures" OR "Risks mitigation" OR "Risks communication" OR "Risks perception" OR "Occupational risk" OR "Chemical risk" OR "Ergonomic risk" OR "Psychosocial risk" OR "Physical risk" OR "Biological risk" OR "Protective measure" OR "Risk mitigation" OR "Risk communication" OR "Risk perception" OR "occupational safety and health" OR "Workers health" OR "Employee Health" OR "Workplace safety" OR "Worker Safety" OR "Safety at work" OR "Professional Risk" OR "Professional Risks" OR "Hazard prediction" OR "work environment" OR "Communication system operations and management" OR "Occupational health" OR "Occupational safety") | 1 | 16:50 |

**Search strategies 11 October 2024**

**Search Strategy for Embase (11 October 2024)**

| Query | Results | Time |
|---|---|---|
| ('health personnel' OR 'personnel, health' OR 'health care providers' OR 'health care provider' OR 'provider, health care' OR 'providers, health care' OR 'healthcare providers' OR 'healthcare provider' OR 'provider, healthcare' OR 'healthcare workers' OR 'healthcare worker' OR 'providers, healthcare' OR 'professional, health care' OR 'health care professional' OR 'health care professionals' OR nurses OR doctors OR 'medical professionals' OR 'healthcare team') AND ('machine learning' OR 'artificial intelligence' OR 'expert system' OR 'cognitive system' OR 'learning, machine' OR algorithms OR 'deep learning' OR 'neural networks' OR 'natural language processing' OR 'cognitive computing' OR 'data mining' OR 'adversarial machine learning' OR boosting OR 'dimensionality reduction' OR 'ensemble learning' OR 'federated learning' OR 'hyperparameter optimization' OR 'multi-armed bandit problem' OR 'random forests' OR 'reinforcement learning' OR 'relevance vector machines' OR 'representation learning' OR 'large language models' OR 'computational and artificial intelligence' OR 'computational intelligence' OR 'feedforward neural networks' OR 'generative adversarial networks' OR 'minimax techniques' OR 'natural languages' OR 'robot learning' OR 'support vector machines' OR 'affective computing' OR 'autonomous robots' OR 'cognitive systems' OR 'decision support systems' OR 'intelligent systems' OR 'knowledge based systems' OR 'learning systems' OR 'prediction methods' OR 'virtual artifact') AND ('risk management' OR 'occupational risks' OR 'chemical risks' OR 'ergonomic risks' OR 'psychosocial risks' OR 'physical risks' OR 'biological risks' OR 'protective measures' OR 'risks mitigation' OR 'risks communication' OR 'risks perception' OR 'occupational risk' OR 'chemical risk' OR 'ergonomic risk' OR 'psychosocial risk' OR 'physical risk' OR 'biological risk' OR 'protective measure' OR 'risk mitigation' OR 'risk communication' OR 'risk perception' OR 'occupational safety and health' OR 'workers health' OR 'employee health' OR 'workplace safety' OR 'worker safety' OR 'safety at work' OR 'professional risk' OR 'professional risks' OR 'hazard prediction' OR 'work environment' OR 'communication system operations and management' OR 'occupational health' OR 'occupational safety') | 246 | 15:55 |

**Search strategies 11 October 2024**

**Search Strategy for Web Of Science (11 October 2024)**

| Query | Results | Time |
|---|---|---|
| ((ALL=("Health Personnel" OR "Personnel, Health" OR "Health Care Providers" OR "Health Care Provider" OR "Provider, Health Care" OR "Providers, Health Care" OR "Healthcare Providers" OR "Healthcare Provider" OR "Provider, Healthcare" OR "Healthcare Workers" OR "Healthcare Worker" OR "Providers, Healthcare" OR "Professional, Health Care" OR "Health Care Professional" OR "Health Care Professionals" OR nurses OR doctors OR "medical professionals" OR "healthcare team")) AND ALL=( "Machine Learning" OR "artificial intelligence" OR "Decision support systems" OR "expert system" OR "Cognitive system" OR "Learning, Machine" OR Algorithms OR "Deep Learning" OR "Neural Networks" OR "Natural Language Processing" OR "Cognitive Computing" OR "Data Mining" OR "Adversarial machine learning" OR Boosting OR "Dimensionality reduction" OR "Ensemble learning" OR "Federated learning" OR "Hyperparameter optimization" OR "Multi-armed bandit problem" OR "Random forests" OR "Reinforcement learning" OR "Relevance vector machines" OR "Representation learning" OR "Robot learning" OR "Large Language Models" OR "Computational and artificial intelligence" OR "Computational intelligence" OR "Feedforward neural networks" OR "Generative adversarial networks" OR "Minimax techniques" OR "Natural languages" OR "Robot learning" OR "Support vector machines" OR "Affective computing" OR "Autonomous robots" OR "Cognitive systems" OR "Decision support systems" OR "Intelligent systems" OR "Knowledge based systems" OR "Learning systems" OR "Prediction methods" OR "Virtual artifact")) AND ALL=("Risk management" OR "Occupational risks" OR "Chemical risks" OR "Ergonomic risks" OR "Psychosocial risks" OR "Physical risks" OR "Biological risks" OR "Protective measures" OR "Risks mitigation" OR "Risks communication" OR "Risks perception" OR "Occupational risk" OR "Chemical risk" OR "Ergonomic risk" OR "Psychosocial risk" OR "Physical risk" OR "Biological risk" OR "Protective measure" OR "Risk mitigation" OR "Risk communication" OR "Risk perception" OR "occupational safety and health" OR "Workers health" OR "Employee Health" OR "Workplace safety" OR "Worker Safety" OR "Safety at work" OR "Professional Risk" OR "Professional Risks" OR "Hazard prediction" OR "work environment" OR "Communication system operations and management" OR "Occupational health" OR "Occupational safety") | 339 | 16:16 |

**Search strategies 11 October 2024**
**Search Strategy for IEEE Xplore (11 October 2024)**

| Query | Results | Time |
|---|---|---|
| ("All Metadata":"Health Personnel" OR "All Metadata":"Personnel, Health" OR "All Metadata":"Health Care Providers" OR "All Metadata":"Health Care Provider" OR "All Metadata":"Provider, Health Care" OR "All Metadata":"Providers, Health Care" OR "All Metadata":"Healthcare Providers" OR "All Metadata":"Healthcare Provider" OR "All Metadata":"Provider, Healthcare" OR "All Metadata":"Healthcare Workers" OR "All Metadata":"Healthcare Worker" OR "All Metadata":"Providers, Healthcare" OR "All Metadata":"Professional, Health Care" OR "All Metadata":"Health Care Professional" OR "All Metadata":"Health Care Professionals" OR "All Metadata":nurses OR "All Metadata":doctors OR "All Metadata":"medical professionals" OR "All Metadata":"healthcare team") AND ("All Metadata":"Machine Learning" OR "All Metadata":"artificial intelligence" OR "All Metadata":"Decision support systems" OR "All Metadata":"expert system" OR "All Metadata":"Cognitive system" OR "All Metadata":"Learning, Machine" OR "All Metadata":Algorithms OR "All Metadata":"Deep Learning" OR "All Metadata":"Neural Networks" OR "All Metadata":"Natural Language Processing" OR "All Metadata":"Cognitive Computing" OR "All Metadata":"Data Mining" OR "All Metadata":"Adversarial machine learning" OR "All Metadata":Boosting OR "All Metadata":"Dimensionality reduction" OR "All Metadata":"Ensemble learning" OR "All Metadata":"Federated learning" OR "All Metadata":"Hyperparameter optimization" OR "All Metadata":"Multi-armed bandit problem" OR "All Metadata":"Random forests" OR "All Metadata":"Reinforcement learning" OR "All Metadata":"Relevance vector machines" OR "All Metadata":"Representation learning" OR "All Metadata":"Robot learning" OR "All Metadata":"Large Language Models" OR "All Metadata":"Computational and artificial intelligence" OR "All Metadata":"Computational intelligence" OR "All Metadata":"Feedforward neural networks" OR "All Metadata":"Generative adversarial networks" OR "All Metadata":"Minimax techniques" OR "All Metadata":"Natural languages" OR "All Metadata":"Robot learning" OR "All Metadata":"Support vector machines" OR "All Metadata":"Affective computing" OR "All Metadata":"Autonomous robots" OR "All Metadata":"Cognitive systems" OR "All Metadata":"Decision support systems" OR "All Metadata":"Intelligent systems" OR "All Metadata":"Knowledge based systems" OR "All Metadata":"Learning systems" OR "All Metadata":"Prediction methods" OR "All Metadata":"Virtual artifact") AND ("All Metadata":"Risk management" OR "All Metadata":"Occupational risks" OR "All Metadata":"Chemical risks" OR "All Metadata":"Ergonomic risks" OR "All Metadata":"Psychosocial risks" OR "All Metadata":"Physical risks" OR "All Metadata":"Biological risks" OR "All Metadata":"Protective measures" OR "All Metadata":"Risks mitigation" OR "All Metadata":"Risks communication" OR "All Metadata":"Risks perception" OR "All Metadata":"Occupational risk" OR "All Metadata":"Chemical risk" OR "All Metadata":"Ergonomic risk" OR "All Metadata":"Psychosocial risk" OR "All Metadata":"Physical risk" OR "All Metadata":"Biological risk" OR "All Metadata":"Protective measure" OR "All Metadata":"Risk mitigation" OR "All Metadata":"Risk communication" OR "All Metadata":"Risk perception" OR "All Metadata":"occupational safety and health" OR "All Metadata":"Workers health" OR "All Metadata":"Employee Health" OR "All Metadata":"Workplace safety" OR "All Metadata":"Worker Safety" OR "All Metadata":"Safety at work" OR "All Metadata":"Professional Risk" OR "All Metadata":"Professional Risks" OR "All Metadata":"Hazard prediction" OR "All Metadata":"work environment" OR "All Metadata":"Communication system operations and management" OR "All Metadata":"Occupational health" OR "All Metadata":"Occupational safety") | 81 | 16:33 |

**Search strategies 11 October 2024**
**Search Strategy for SCOPUS (11 October 2024)**

| Query | Results | Time |
|---|---|---|
| ( TITLE-ABS-KEY ( "Health Personnel" OR "Personnel, Health" OR "Health Care Providers" OR "Health Care Provider" OR "Provider, Health Care" OR "Providers, Health Care" OR "Healthcare Providers" OR "Healthcare Provider" OR "Provider, Healthcare" OR "Healthcare Workers" OR "Healthcare Worker" OR "Providers, Healthcare" OR "Professional, Health Care" OR "Health Care Professional" OR "Health Care Professionals" OR nurses OR doctors OR "medical professionals" OR "healthcare team" ) AND TITLE-ABS-KEY ( "Machine Learning" OR "artificial intelligence" OR "Decision support systems" OR "expert system" OR "Cognitive system" OR "Learning, Machine" OR algorithms OR "Deep Learning" OR "Neural Networks" OR "Natural Language Processing" OR "Cognitive Computing" OR "Data Mining" OR "Adversarial machine learning" OR boosting OR "Dimensionality reduction" OR "Ensemble learning" OR "Federated learning" OR "Hyperparameter optimization" OR "Multi-armed bandit problem" OR "Random forests" OR "Reinforcement learning" OR "Relevance vector machines" OR "Representation learning" OR "Robot learning" OR "Large Language Models" OR "Computational and artificial intelligence" OR "Computational intelligence" OR "Feedforward neural networks" OR "Generative adversarial networks" OR "Minimax techniques" OR "Natural languages" OR "Robot learning" OR "Support vector machines" OR "Affective computing" OR "Autonomous robots" OR "Cognitive systems" OR "Decision support systems" OR "Intelligent systems" OR "Knowledge based systems" OR "Learning systems" OR "Prediction methods" OR "Virtual artifact" ) AND TITLE-ABS-KEY ( "Risk management" OR "Occupational risks" OR "Chemical risks" OR "Ergonomic risks" OR "Psychosocial risks" OR "Physical risks" OR "Biological risks" OR "Protective measures" OR "Risks mitigation" OR "Risks communication" OR "Risks perception" OR "Occupational risk" OR "Chemical risk" OR "Ergonomic risk" OR "Psychosocial risk" OR "Physical risk" OR "Biological risk" OR "Protective measure" OR "Risk mitigation" OR "Risk communication" OR "Risk perception" OR "occupational safety and health" OR "Workers health" OR "Employee Health" OR "Workplace safety" OR "Worker Safety" OR "Safety at work" OR "Professional Risk" OR "Professional Risks" OR "Hazard prediction" OR "work environment" OR "Communication system operations and management" OR "Occupational health" OR "Occupational safety" ) ) | 718 | 16:22 |
